# Annotating off-label drug usage from unconventional sources

**DOI:** 10.1101/2022.09.08.22279709

**Authors:** Sorin Avram, Liliana Halip, Ramona Curpan, Ana Borota, Alina Bora, Tudor I Oprea

**Affiliations:** Department of Computational Chemistry, “Coriolan Dragulescu” Institute of Chemistry Timisoara of the Romanian Academy, 24th Mihai Viteazu Av., 300223 Timisoara, Romania; Roivant Discovery Sciences, Boston, MA, USA

**Author notes:** **Corresponding author:** Tudor I. Oprea, Roivant Discovery Sciences, Boston, MA 02210, USA.

**Keywords:** off-label, drug indication, clinical trial, drug prescription, data mining

## Abstract

Physicians have the freedom to prescribe medicines outside the list of approved indications, to treat mild to life-threatening clinical conditions and diseases, particularly when conventional treatments fail or are lacking. Off-label drug usage is more frequently observed in specific populations not often represented in clinical trials, e.g., pediatric, geriatric, or pregnant patients. Despite conflicting reports on patient safety, exploring alternative treatment options in medical practice promotes innovation and extends the applicability of current medicines. This process can be significantly improved by properly documenting and discussing off-label usage. This paper aims to document off-label uses discussed in less conventional sources, such as the r/medicine Reddit subforum. We identified 66 “Reddit off-label uses” (ROLUs) not captured in our reference database, DrugCentral (https://drugcentral.org/), for a set of 40 drugs. These drugs are associated with 209 on-label drug indications (INDs) and 58 “non-Reddit” off-label uses (NROLUs). Most of these drugs are relatively ‘old’ (approved before 2000) and act on the nervous system, many with psychiatric applications. However, ROLUs are distinct from INDs and NROLUs. An automated scientific literature query showed that 90% of the ROLUs are linked to 4 scientific publications or more, with 80% linked to at least 10. A further search in the clinical trials database revealed 46 ROLUs mentioned; 39 are in phase 3 trials. These results indicate that most off-label uses discussed on the Reddit forum are supported by scientific evidence. We conclude that medical social media channels can provide a valuable source of alternative drug applications and should be scientifically explored and evaluated in the future.

## INTRODUCTION

Off-label drug use refers to the prescription of marketed medications not approved by regulatory agencies such as the U.S. Food and Drug Administration (for the United States) and not listed in the required drug-labeling information.^1^ Physicians may choose to treat mild to life-threatening clinical conditions and diseases outside the approved uses for patients that respond poorly to conventional treatment, often in specific populations less likely to be included in clinical trials (e.g., pediatric, pregnant, or psychiatric patients).^1^ If two diseases share similar pathological or physiological mechanisms, a physician may use a drug approved for the first disease to treat the second condition off-label (e.g., anxiety and posttraumatic stress disorder).^1^ Changes in pharmaceutical formulations (e.g., oral solution instead of capsules) or dosage (e.g., two tablets instead of one per day) for an indication is also considered off-label drug use.^2^

Off-label drug use lacks rigorous regulations.^3^ Despite conflicting reports on patient safety,^4,5^ it is a common and legal practice. A literature survey^6^ found that off-label uses (1985–2004) ranged between 11%–80% of all drug uses,^6,7^ with higher rates in pediatric patients^8^ versus adults^9^ and in inpatient vs. outpatient settings.^6^

Despite its relevance, scientific support for off-label uses is relatively poor.^7^ However, the freedom granted to physicians to explore alternative drug uses has been shown to promote innovation and extend current therapeutic applications.^10^ One study found that 57% of drug therapy innovations were discovered by practicing clinicians through field discovery.^10^ To accelerate such innovation, off-label uses should be documented and openly discussed.

Some digital resources attempt to mine off-label drug uses^11,12^ from conventional sources of drug uses, such as scientific literature (articles, case studies, reviews)^12^ or the US National disease and therapeutic index (NDTI), where office-based physicians share information on prescribing patterns and treatments of diseases.^7^

Here, we document data extraction of off-label uses from a less conventional data source, the r/medicine Reddit subforum (Meddit).^13^ Following data extraction, we performed an automated search in the biomedical literature to assess the scientific validity of these medical uses and compared the results to on-label indications. In addition, we queried clinical trials for drug use and summarized the results.

## MATERIALS AND METHODS

### Data retrieval

In April 2021, we manually extracted potential off-label uses from meddit - *“What are unconventional, off-label uses of common medications in your specialty?”*^*13*^ The drugs were mapped onto DrugCentral^11^ (accessed October 2021), and the corresponding off-label uses were examined. We found 66 drug uses: Fifty-one unique clinical conditions/diseases associated with 40 drugs were not captured in DrugCentral 2021. This dataset is designated as the ROLU (Reddit off-label uses) data set for the remainder of the text. For the 40 drugs covered by Reddit data, we retrieved 209 indications (further referred to as the IND data set) and 58 non-Reddit off-label uses (the NROLU data set) from DrugCentral 2021.

### MedDRA ontology mapping

All medical terms that define drug uses (indications and off-label uses) were mapped into the MedDRA dictionary. MedDRA, the Medical Dictionary for Regulatory Activities, is the international medical terminology developed under the auspices of the International Council for Harmonization of Technical Requirements for Pharmaceuticals for Human Use (ICH). The dictionary holds a medical dictionary designed hierarchically from specific medical conditions and diseases up to general system organ classes. Low-level terms (LLTs) define closely related medical terms (synonyms) grouped into preferred terms (PTs). Further, PTs are grouped into more general high-level terms (HLTs), further into high-group level terms (HGLTs), and finally in system organ classes (SOCs). In MedDRA, a PT can be shared by two or multiple superior hierarchical levels: HLT, HGLTs, or SOCs. For example, “migraine” (PT) is shared by two HLTs “migraine headaches” (found in nervous system disorders SOC, “Nerv”) and “Cerebrovascular and spinal vascular disorders NEC” (found in vascular disorders SOC, “Vasc”).

We used LLTs, PTs, and HLTs to map diseases and conditions describing drug indications and off-label uses in DrugCentral, Meddit, scientific literature, and clinical trials (*vide infra*) data. The procedure, fully automated via the R statistical programming platform,^14^ maps these terms in scientific literature and clinical trials. We manually assigned PTs for DrugCentral uses and ROLU data.

### Clinical Trials data retrieval

Clinical trials data was downloaded in XML format (N = 311,342 clinical trials, or CTs) from the International Clinical Trials Registry Platform (ICTRP)^15^ (accessed November 5, 2021) maintained by the World Health Organization (WHO). First, we used drug synonyms from DrugCentral to map the 40 drugs in the “Intervention” section. Out of the 8,509 processed CTs, we matched at least one disease/medical condition (drug uses and corresponding MedDRA terms) in 7,093 (83%) of the entries. After querying the “Condition” section of the clinical trials, we grouped this dataset into 6967 drug-PT pairs (herein termed the CT dataset), then tallied the number of CTs associated with each drug.

### PubMed data retrieval

PubMed records were retrieved from NCBI (National Center for Biotechnology Information)^16^ using the “easyPubmed” R package.^14^ We built an advanced search syntax querying the name of the drugs and the medical term defining the clinical conditions and diseases for drug uses in ROLU, IND, and NROLU. Unique PubMed identifiers (PMIDs) were retrieved and grouped for 333 drug uses. In addition, according to the data provided by PubMed, for each drug use, we tallied the number of case reports, CTs (as the sum of randomized and controlled CTs), and meta-analysis papers.^17^

## RESULTS AND DISCUSSIONS

### Description of ROLU drugs

The ROLU set has 66 off-label uses for 40 drugs (Tabel 1) extracted from Meddit (Data_sets.xlsx - Supporting Information). According to our classification scheme based on proprietary rights and marketing status implemented in DrugCentral,^18^ these drugs are “off-patent.” Except for ezetimibe, duloxetine, bevacizumab, and capsaicin, the other 36 drugs (90%) were first FDA approved before 2000, with 25 (62.5%) having been FDA approved before 1990 (Table S1 in SI.pdf - Supporting Information).

More than half (58%) of these drugs act on the nervous system, covering psychiatric applications (see Figure 1), with mechanism-of-actions interfering with GABA, cholinergic, serotoninergic, dopaminergic, noradrenergic, or histaminergic pathways. These drugs are antipsychotics (olanzapine, haloperidol, prochlorperazine), antidepressants (mirtazapine, duloxetine, doxepin, sertraline), hypnotics (zolpidem, phenobarbital), antiepileptic (phenytoin, phenobarbital, gabapentin), antiparkinsonians (levodopa, amantadine), or antihistamines (cyproheptadine, loratadine, diphenhydramine, fexofenadine), respectively. Some drugs extend their on-label uses as antiemetics and muscle relaxants (Table S1 in SI.pdf - Supporting Information). Other representatives include insulin (antidiabetic), allopurinol (antigout agent), ezetimibe and simvastatin (antihyperlipidemic agents), and alendronic acid (bone resorption inhibitor), followed by cardiovascular drugs, e.g., diltiazem and phenytoin (antiarrhythmics), pentoxifylline and nimodipine (vasodilators), etc. – see Figure 1. Apart from drug indications, DrugCentral provided 58 off-label uses (NROLU) for 25 of the 40 drugs (Table 1). Here, the top five drug application areas (53% of the drug uses) concern the nervous system, psychiatric, muscular (and connective tissue), immune system disorders, and injuries and poisonings.

**Figure 1.**
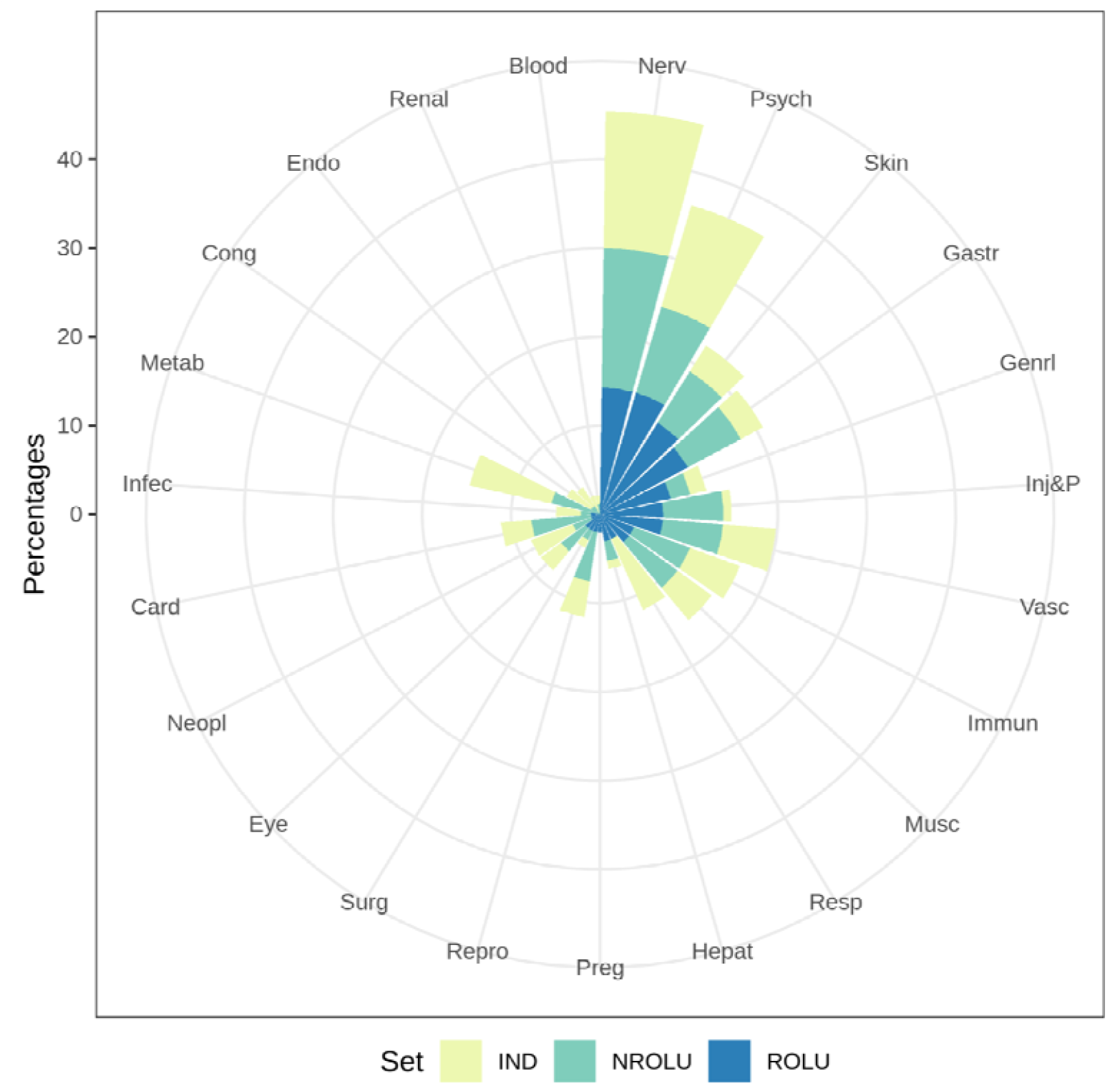
Percentages of therapeutic uses (MedDRA SOC) within ROLU, IND, and NROLU data sets.

**Table 1.** Summary of the three data sets.

| Data set | Num. of drugs | Num. of drug uses<sup>1</sup> | Num of drug uses |  |
| --- | --- | --- | --- | --- |
|  |  |  | Mapped as PT/HLT/SOC terms<sup>2</sup> | Clinical Trials<sup>3</sup> |
| ROLU | 40 | 66 | 65/94/86 | 47 |
| IND | 40 | 209 | 185/200/130 | 154 |
| NROLU | 25 | 58 | 56/72/58 | 44 |

<sup>1</sup> drug-indication/off-label use pairs (drug use) according to DrugCentral (IND, NROLU) and to Reddit (ROLU); <sup>2</sup> MedDRA - preferred terms (PTs), higher-level terms (HLT) and System Organ Classes (SOC) drug-uses; <sup>3</sup> drug uses retrieved from WHO clinical trials database.

### Drug use comparison between ROLU, IND, and NROLU data

There is little overlap between ROLU, IND, and NROLU (see Figure 2A). However, we identified closely related uses between these sets by examining the higher level of MedDRA hierarchy (HLT). For example, doxepin, a histamine H1 and H2-receptor blocker with psychotropic activity, is indicated for pruritus (skin itch) in topical cream formulations but used off-label for *cholestatic* pruritus according to ROLU (Figure 2B). Another example of HLT-based ROLU/NROLU similarity (Table S2 in SI.pdf - Supporting Information) is the use of gabapentin for chronic pain (ROLU) and acute postoperative pain (NROLU; Figure 2B).

**Figure 2.**
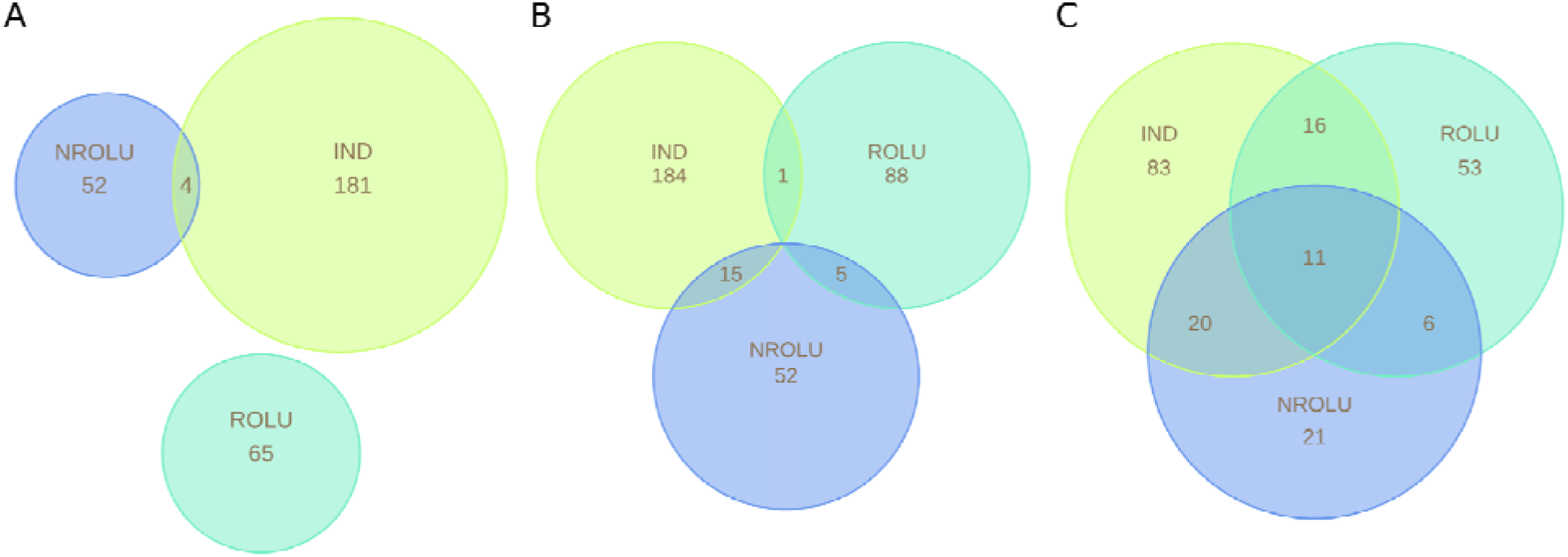
Venn diagram showing the overlap between drug uses based on MedDRA PT (A), HLT (B), and SOC (C) in ROLU, IND, and NROLU sets.

Most ROLU are primarily associated with nervous system and psychiatric pathologies (38%), with 43% of ROLUs (for 17 drugs) being similar to IND uses, followed by skin (and subcutaneous), gastro-intestinal and general disorders respectively (Figure 1).

At the therapeutic level (SOC, Figure 2C), there are 11 cases of drug uses shared between the three sets. For example, gabapentin is used in three disorders related to the nervous system: insomnia (ROLU), postherpetic neuralgia (IND), and essential tremor (NROLU).

### Scientific literature support for drug uses

Scientific evidence was estimated based on the number of publications indexed in PubMed (PMIDs) co-mentioning the drug and associated disease in the title or abstract. The results indicate a median of 36 PMIDs/off-label use, with 90% of the ROLU entries linked to ≥ 4 PMIDs, and 80% to ≥ 10 PIMDs (Figure 3A).

**Figure 3.**
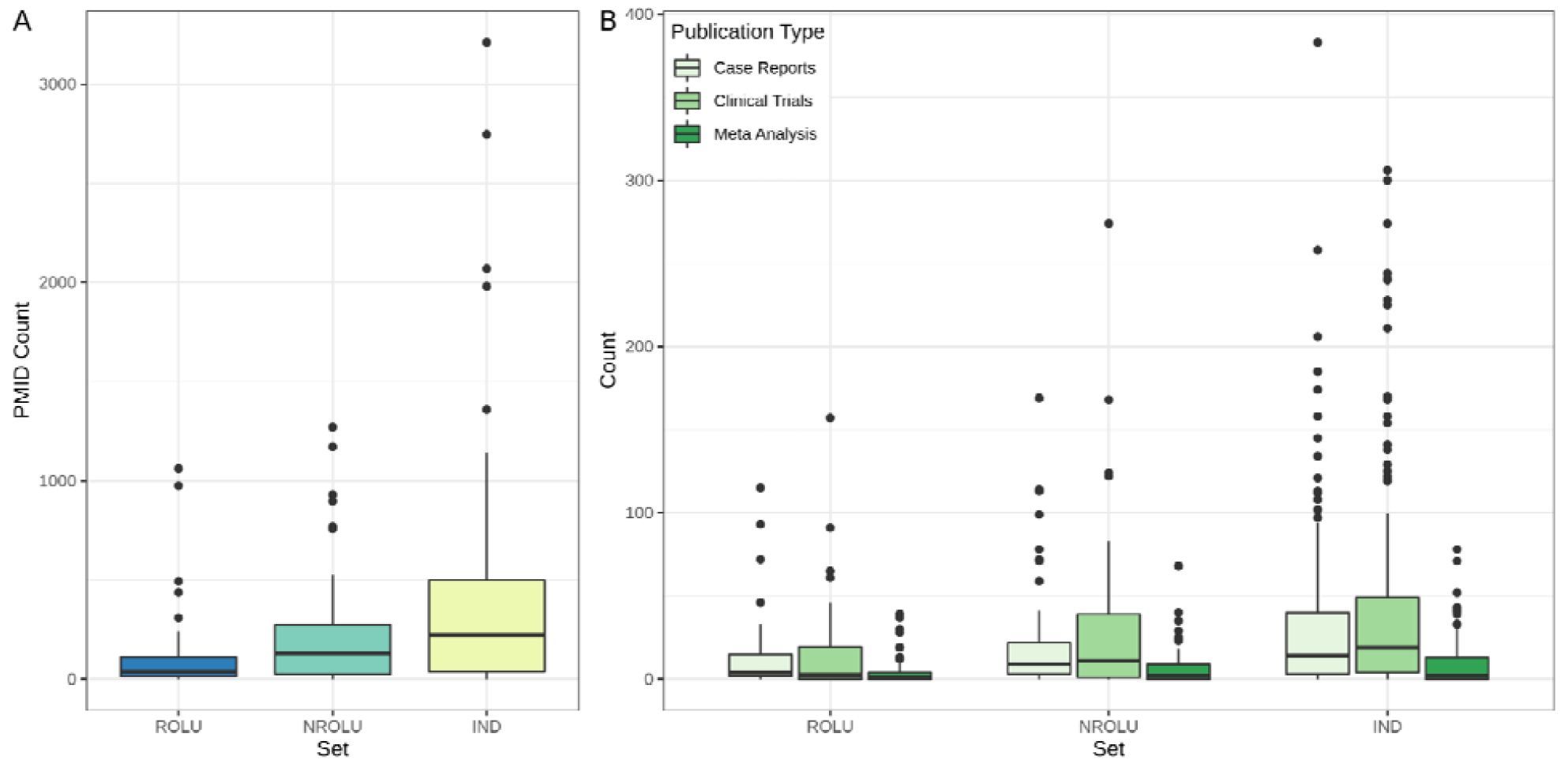
Boxplots of the number of publications found in PubMed (Pubmed IDs - PMIDs) for drug applications (covered by 40 drugs extracted from Reddit) comprised in IND (DrugCentral indications), NROLU (non-Reddit off-label uses DrugCentral), and ROLU (Reddit off-label uses).

With over 900 publications, the most frequently mentioned off-label uses refer to gabapentin (approved for postherpetic neuralgia) and ketamine (anesthetic agent) for pain treatment (Table 2). The results on gabapentin are consistent with observations^7^ from Radley et al. package one of the most studied off-label use.

**Table 2.** Examples of ROLU with extreme numbers of PMID mappings.

| Num of PMIDs<sup>1</sup> | Drug name | Drug class | Primary indication | Reddit off-label use |
| --- | --- | --- | --- | --- |
| >900 | gabapentin | Gabamimetic agent | Postherpetic neuralgia | Chronic pain |
|  | ketamine | Anesthetic agent | Anesthesia | Pain |
| $\leq 1$ | dicycloverine | Spasmolytic with a papaverine-like action agent | Irritable bowel syndrome | Calculus in biliary tract |
|  | protamine sulfate | Heparin reversal agent | Heparin overdose | Neuropathic pain |
|  | levocarnitine | Amino acid derivatives | Carnitine deficiency | Recovery during viral myocarditis |
|  | alendronic acid | Bisphosphonates | Osteoporosis | Complex regional pain syndrome type I |

<sup>1</sup> PubMed IDs (https://pubmed.ncbi.nlm.nih.gov/)

In contrast to heavily discussed drugs uses, we identified 4 cases with one or no supporting publications (Table 2), which is not unusual for off-label uses.^7^ However, supporting evidence could be sought for closely related drugs evaluated for the same use. For example, dicycloverine is used for treating cholelithiasis (i.e., calculus in the biliary tract) *cf*. ROLU. By altering the search to “spasmolytic” and “cholecystitis,” we found several publications. One report describes the benefic effects of mebeverine (a papaverine-like spasmolytic similar to dicycloverine) in cholecystitis.^19^ Another report reviewed multiple CTs and found contradicting results on whether NSAIDs or spasmolytics (scopolamine, hyoscine) are better suited for pain management in cholelithiasis.^20^ No publication supports the use of protamine sulfate (approved as a heparin reversal agent) in neuropathic pain (Table 2). However, deriving the medical term to the more general “pain” allowed us to identify some evidence supporting this off-label use.^21^ Study results for the use of levocarnitine in recovery during viral myocarditis^22^ and for alendronic acid to treat complex regional pain syndrome^23^ type I are promising but limited.

Regarding publication type (Figure 3B), the ROLU are primarily supported by case reports (80%) and clinical trials (70%). Meta-analyses were found for half of the ROLU. For the IND and NROLU sets, the number of associated publications is larger (for all types of publications), particularly for indications (a 6-fold larger median, as shown in Figure 3A). Most of the on-label drug uses (85%) are linked to at least one case report and clinical trial, and 65% to meta-analyses papers.

### Off-label uses in clinical trial data

To consolidate our assessment of the scientific support for ROLU, we explored the WHO’s ICTRP database of clinical trials. We retrieved 6277 CTs matching 71-76% of drug uses in ROLU, NROLU, and IND sets (Table 1). For the majority of the drug uses (80%), we found up to 9 (median of 4), 12 (median of 5), and 32 (median of 8) CTs in ROLU, NROLU, and IND, respectively (Data_sets.xlsx - Supporting Information). Consistent with the literature search results, using ketamine and gabapentin for pain management (extracted from Reddit) is supported by the largest CT numbers (135 and 60, respectively). We found that 46 ROLU uses are mentioned in phase 2 CTs. These results support the literature search outcomes and show scientific support for 82% (39 of 46 uses are also in phase 3 CTs) of the Reddit drug uses found in CTs.

### Limitations

The current study suffers from several limitations. Firstly, drug uses revolve around medical terms and ontology-based studies that are subject to terminology variations. For example, the number of unique drug indications dropped from 209 to 185 when mapped to MedDRA preferred terms (Table 1). Some on-label uses as indexed in DrugCentral are specific, whereas MedDRA terms are more general. MedDRA mappings are provided as supplementary data (Data_sets.xlsx - Supporting Information).

A second major limitation might be caused by the age of the drugs and the limited availability of electronic documentation. Many are older drugs (50% were approved before 1990, see Table S1 in SI.pdf - Supporting Information), and supporting evidence (PubMed and CTs) might not be consistently described electronically. For example, the use of spasmolytics for pain control in cholelithiasis, discussed above, is supported by CTs from the 1980s and 1990s.^20^

Thirdly, heterogeneity in clinical trial description might also limit the results. For example, only 70% of the drug indications could be matched in clinical trial data. Another aspect concerns the concluding statements from publications and the CT outcomes: These were not individually analyzed, given the large volume of data. As mentioned above, the current study concerns off-label uses where the clinical condition/disease differs from the drug indication. However, we did not analyze off-label uses that require changes in posology, formulation, etc.

## CONCLUSION

Because most of the drugs investigated herein interfere with neurotransmitters, their approved and off-label uses address diseases related to the nervous system and psychiatric disorders. Social media posts such as Meddit discuss off-label uses that differ from drug indications and well-annotated off-label uses, as captured in publicly available pharmaceutical databases. Meddit posts are less scientifically documented, partly due to their informal style, but are more likely to capture novelty since medical practitioners wrote them. However, we found supporting evidence for therapeutic efficiency for most of the ROLUs in literature and CT data. These results suggest a benefit in surveying Meddit posts to enhance pharmaceutical knowledge regarding novel off-label uses. The 2023 release of DrugCentral will capture these ROLUs, in order to enhance the coverage of therapeutic drug uses for the 40 drugs discussed here. We conclude that medical social media channels can provide a valuable source of alternative drug applications and should be scientifically explored and evaluated in the future.

## Data Availability

All data produced in the present work are contained in the manuscript

## SUPPORTING INFORMATION

Supplementary information accompanies this paper: “Data_sets.xlsx” and “SI.pdf”.

## ACKNOWLEDGMENTS

MedDRA® trademark is registered by ICH.

