## Supplemental Material 1 for "Annotating off-label drug usage from unconventional sources"

Table S1. A brief description of the drugs in Reddit off-label use set (ROLU)............................ 2

Table S2. Overlapping off-label uses in ROLU and NROLU sets based on MedDRA's HLT mapping. ........................................................................................................................ 8

**Table S1.** A brief description of the drugs in Reddit off-label use set (ROLU).

| DrugCentral ID | Drug name | FDA first approval year | Drug Class |
|---|---|---|---|
| 112 | alendronic acid | 1995 | bisphosphonate derivatives; calcium metabolism modifiers |
| 124 | allopurinol | 1966 | Purine derivative; xanthine oxidase inhibitor; antigout agent |
| 144 | amantadine | 1968 | tricyclic compounds; histamine-H1 receptor antagonists, dopamine agonists; antiparkinson agent; antivirals |
| 282 | baclofen | 1977 | GABA derivative; GABA-B receptor agonist; skeletal muscle relaxant agent |
| 513 | levocarnitine | 1985 | amino acid derivative; essential cofactor for fatty acid metabolism; muscle deficiencies |
| 765 | cyproheptadine | 1961 | tricyclic compounds, histaminic-H1 receptor and serotonin antagonists; anti-allergic agent |
| 842 | dextromethorphan | 1957 | morphine derivate; opioid receptor antagonists/agonists; antitussive |

| | | | |
|---|---|---|---|
| 868 | dicycloverine | 1950 | muscarinic acetylcholine receptor antagonist; spasmolytics with a papaverine-like action |
| 897 | diltiazem | 1982 | benzothiazepine derivative, calcium channel blockers, Antidysrhythmic IV agent; |
| 916 | diphenhydramine | 1946 | aminoalkyl ether derivatives, histamine-H1 receptor antagonist, antihistamines; antiemetic, antiallergic and sedative agents |
| 956 | doxepin | 1969 | tricyclic compounds, dibenzoxepin; non-selective monoamine reuptake inhibitors, anticholinergic activity and histamine H1/2 receptors antagonists; antidepressant agent |
| 972 | duloxetine | 2004 | thiophene derivative; serotonin and/or norepinephrine reuptake inhibitors, fluoxetine derivatives; antidepressants |
| 1125 | ezetimibe | 2002 | 2-azetidinones derivative; acyl CoA: cholesterol acyltransferase (ACAT) inhibitors; antihyperlipidaemic agent |

| | | | |
|---|---|---|---|
| 1170 | fexofenadine | 1996 | Piperidine dereiviative, metabolite of terfenadine; histamine H1 receptor antagonists; second generation antihistamine antiallergic agent |
| 1171 | finasteride | 1992 | azasteroid antiandrogen; testosterone-5-alpha reductase inhibitors; antihyperplastic agent |
| 1264 | gabapentin | 1993 | GABA analog; gabamimetic agent; analgesic and antiepileptic agent |
| 1353 | haloperidol | 1967 | phenyl-piperidinyl-butyrophenone derivative; 1st Generation antipsychotics |
| 1523 | ketamine | 1970 | cyclohexanone derivative, N-methyl-D-aspartate (NMDA) receptors inhibitor; anesthetic agent |
| 1567 | levodopa | 1970 | dopamine derivatives, dopamine receptor agonists; antiparkinsonism/prolactin inhibitors |
| 1605 | loratadine | 1993 | tricyclic compounds, histaminic-H1 receptor antagonists; second generation antihistaminic, antiallergic agent |

| | | | |
|---|---|---|---|
| 1816 | mirtazapine | 1996 | piperazinoazepine tetracyclic compounds; alpha-2 adrenergic, 5-HT2/3 serotonin receptors, histamine H1 receptor anatagonist, antidepressants |
| 1937 | nimodipine | 1988 | phenylpyridine (nifedipine) derivatives; calcium channel blockers; vasodilator cardiovascular agent |
| 1982 | olanzapine | 1996 | benzodiazepine tracyclic compounds; serotonin , histamin, alpha-1 adrenergic, dopamine and muscarine recepor modulator; 2nd Generation, antimanic agents |
| 2099 | pentoxifylline | 1984 | N-methylated xanthine derivatives (purine derivative); phosphodiesterase inhibitor; vasodilator and antiaggregant agents |
| 2134 | phenobarbital | 1912 | barbituric acid derivatives; GABA modulators; antiepileptic, anticonvulsant , hypnotic, sedative agent |
| 2152 | phenytoin | 1953 | hydantoin derivatives; sodium channel blockers; antiepileptic, anticonvulsants, antiarrhythmic and muscle relaxant agent |

| | | | |
|---|---|---|---|
| 2274 | prochlorperazine | 1956 | phenothiazine derivative; alpha-adrenoceptor and dopamine receptor antagonist; antiemetic and antipsychotic agent, |
| 2436 | sertraline | 1991 | 1-naphthylamine derivative; selective serotonin reuptake inhibitor (SSRI); antidressant agent |
| 2445 | simvastatin | 1991 | lovastatin derivative; HMG CoA reductase inhibitors;statins, antihyperlipidaemic agent |
| 2475 | spironolactone | 1960 | steroidal antiandrogenic compound; antimineralocorticoid, selective aldosterone antagonists; potassium-sparing diuretics |
| 2561 | tamoxifen | 1977 | stilbene derivative; selective estrogen receptor antagonists; antineoplastic agent |
| 2616 | thalidomide | 1998 | piperidinyl isoindole derivative, tumor necrosis factor inhibitor, angiogenesis inhibitor; antiinflammator, cytostatic and immunosuppressant agent |
| 2713 | tranexamic acid | 1986 | antifibrinolytic, hemostatic agent |

| | | | |
|---|---|---|---|
| 2870 | zolpidem | 1992 | imidazopyridine derivative and short-acting GABA-A receptor agonist; hypnotics/sedatives agent |
| 3064 | capsaicin | 2009 | alkylamide; transient receptor potential cation channel blocker; anesthetic agent |
| 4061 | timolol | 1978 | beta-blockers (propranolol type) |
| 4215 | isopropanol | | isomer of 1-propanol; topical antispetic |
| 4956 | bevacizumab | 2004 | monoclonal antibodies of humanized origin, VEGF Inhibitor; Antineoplastics |
| 5003 | insulin human | 1982 | hormone; rapid-acting insulins; antidiabetics |
| 5034 | protamine sulfate | 1969 | proteins; anticoagulant neutralizing agent |

**Table S2.** Overlapping off-label uses in ROLU and NROLU sets based on MedDRA's HLT mapping.

| Drug_name | HLT name | ROLU Drug Use | NROLU Drug Use |
|---|---|---|---|
| gabapentin | Pain and discomfort | Chronic pain | Acute postoperative pain |
| amantadine | Cerebral injuries | Arousal post traumatic brain injury | Cognitive Impairment following Traumatic Brain Injury |
| haloperidol | Nausea and vomiting symptoms | Nausea and vomiting | Chemotherapy-induced nausea and vomiting |
| olanzapine | Nausea and vomiting symptoms | Nausea and vomiting | |
| | | cannabinoid hyperememsis | |
| tranexamic acid | Haemorrhages | Epistaxis | Hyperfibrinolysis Induced Hemorrhage |
| | | | Postsurgical Hemorrhage |
